# Vascular risk factors modulate gender-specific aging of brain white matter structural network

**DOI:** 10.1101/2023.06.15.23291436

**Authors:** Haojie Chen, Weijie Huang, Xinyi Dong, Guozheng Feng, Yiwen Wang, Zhenzhao Liu, Shuning Ma, Junjie Peng, Li Su, Ni Shu

## Abstract

Vascular risk factors (VRFs) are known to increase risk for cerebrovascular disease and dementia, such as Alzheimer’s disease that is described as a dysconnectivity syndrome. The gender-related evidence on associations between VRFs and white matter (WM) structural network in large community-dwelling populations across middle and older age will contribute to understanding the biological underpinnings of sex and gender considerations in dementia. Based on 17,954 participants from the UK Biobank, we present the relationship between VRFs and WM network architecture (measured with network integration and segregation) in different gender groups. First, females exhibit lower network architecture and experience an accelerated decline earlier than males. Second, network integration is more sensitive to VRFs than segregation, with diabetes, hypertension, and excessive alcohol consumption having the greatest impact. Third, we found greater susceptibility of network architecture to VRFs in males, as well as female-preferred effects on regional integration of obesity, particularly on the subcortical structure and occipital lobe. Finally, higher combined risk was associated with more disrupted network architecture particularly on temporal and frontal lobe, as well as lower processing speed and working memory in both genders. Our findings provide new insights into understanding the relationship between VRFs and WM network architecture, guiding interventions to promote successfully cognitive aging and highlighting the importance of considering gender-specific effects in future research.

## Introduction

The age-related cognitive impairment is common in the increasingly ageing population. It is of great importance to understand the neurobiological underpinnings of the impairment. The proposal of “human brain connectome” provides a new perspective to study the biological mechanisms underlying the age-related cognitive impairment^1^. A plethora of evidence suggest that neurodegenerative disease can be regarded as “disconnection syndrome” and the cognitive impairment is contributed to a loss of neurons and their connections that will interfere with the white matter (WM) connectivity between macroscopic brain regions^2^. Discriminating the modifiable risk factors leading to impairment of brain connectome and decline of cognition is urgently needed, as modification of these risk factors can be a cost-effective preventive strategy for improving brain health.

Emerging evidence suggests that brain health and cardiovascular health have shared risk factors^3^ and controlling vascular risk factors (VRFs) may delay the cognitive decline and even reduce the incidence of dementia^4–6^. These VRFs, including vascular diseases such as hypertension and diabetes, as well as unfavorable lifestyles such as excessive alcohol consumption, smoking, and obesity, have consistently been found detrimental to the brain structure and function^5^. Although prior studies have investigated the effects of VRFs on the architecture of the WM network^7–10^, most studies have focused on only a single VRF, and their results varied considerably due to small sample sizes and heterogeneity. For example, a recent study found no significant association between obesity and aging WM network architecture^11^, but Rahmani and colleagues identified gender-specific effects of increasing body mass index on WM network^12^. Therefore, large, homogeneous cohorts with comprehensive surveys are necessary to compare the effect sizes and the patterns of impairment caused by VRFs on the WM network, particularly with regards to gender-related effects.

Gender differences in the aging brain have garnered significant attention as aging females typically exhibit worse performance in spatial tasks and higher performance in verbal tasks^13^, as well as higher prevalence of neuropsychological disorders^14^. Neuroimaging studies have provided insight into the underlying biological mechanisms that contribute to these gender differences: Males demonstrate greater raw white matter and gray matter volumes, WM integrity, and functional connectivity in the sensorimotor network and default mode network throughout the aging process^15^. However, there remains a paucity of knowledge regarding gender differences in the aging WM wiring topology that underlies between-neuronal communication^16^. Despite gender has been considered as a covariate in large-scale human connectome studies, gender effects on aging WM network architecture have not been investigated to the same extent as volumetric measures. Therefore, elucidating the gender difference in WM architecture within the context of aging and VRFs may unveil novel insights into the neural foundations of disconnection syndromes^17^.

To investigate the effect of VRFs on gender-specific aging of WM network, we analyzed the integration and segregation of 17,954 WM connectomes from UK Biobank. UK Biobank recruited 500,000 people aged between 40 - 69 years in 2006 - 2010 from across the UK, with detailed information about lifestyle, physical measures, and blood chemistry analyses. Our study aims to address the following questions: (i) Does the WM network architecture demonstrate sensitivity to aging and gender comparable to volumetric measures? (ii) What are the gender-common and gender-specific WM network impairments result from individual VRF? (iii) How does the combined effect of VRFs on WM network architecture and cognition impact males and females?

## Results

### Population characteristics

In this study, we analyzed a cohort of 17,954 cognitively normal individuals with available Magnetic Resonance Imaging (MRI) scans and no history of neuropsychiatric diseases, selected from a pool of over 500,000 participants in the UK Biobank project. Significant gender differences were observed in all demographic characteristics, brain imaging measures, and cognitive test performances (Figure 1a, all P<0.01 except for Apolipoprotein E (APOE) ε4 carrier proportion). Male participants were on average older (63.9 ± 7.54 years vs. 62.5 ± 7.28 years) and have a higher ratio of college qualification (47.8% vs. 45.0%) than females. We considered five VRFs, consisting of two common vascular diseases (diabetes and hypertension) and three unfavorable lifestyle factors (smoking, excessive alcohol consumption, and obesity) (Figure 1a, see **Supplementary Table 1** for definition of each VRF). The proportion of individuals who reported being alcoholic was lower among males than females, while males had a higher proportion of smokers and obese individuals (Figure 1a, see **Supplementary Table 2** for demographic statistics). Males also had higher ratios of hypertension and diabetes than females. Exclusion criteria and VRFs definitions are detailed in the Methods section.

**Figure 1.**
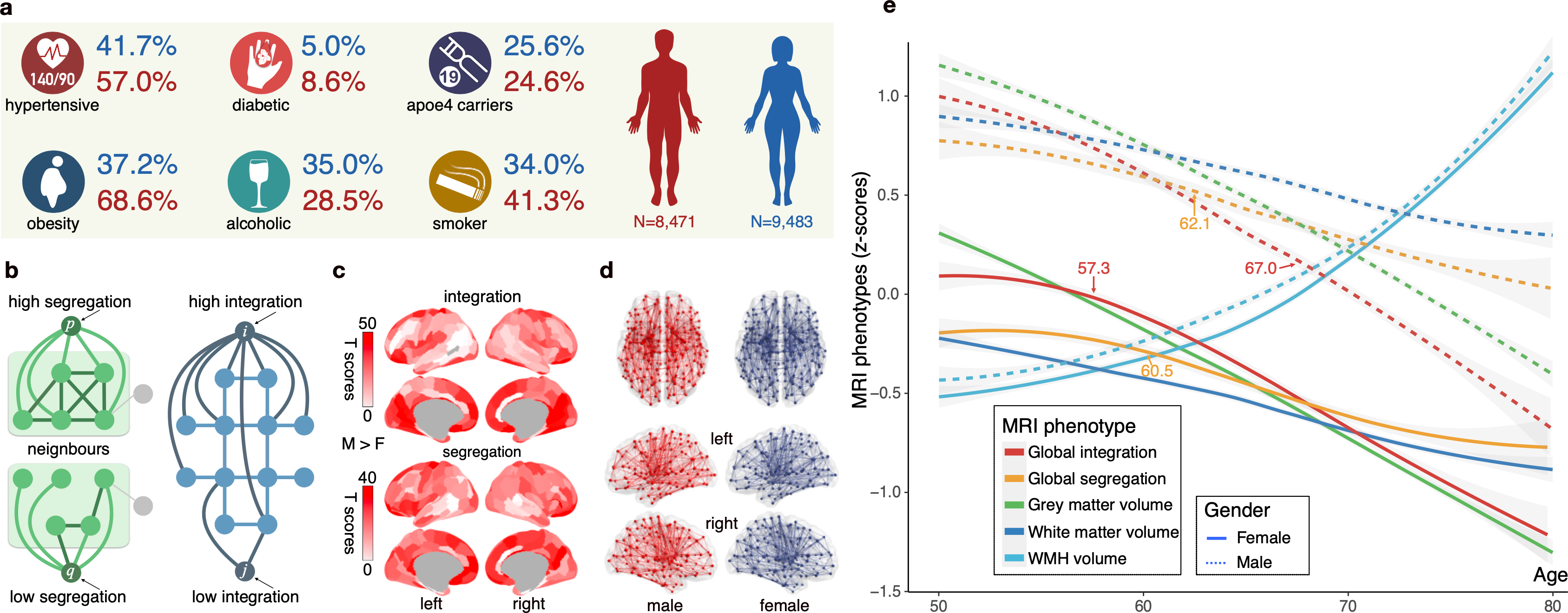
| Population Characteristics and Aging Trajectories of MRI Phenotypes. **a**, Population Characteristics. This figure includes data from UK Biobank, comprising 8,741 males (shown in red) and 9,483 females (shown in blue). We considered VRFs including diabetes, hypertension, excessive alcohol consumption, smoking, obesity as well as APOE ε4 carrier status. Among the population, the proportion of individuals reporting alcoholism was lower among males than females (28.5% vs. 35.0%). Men had a higher proportion of smokers (41.3% vs. 34.0%) and obese individuals (68.6% vs. 37.2%). Males also had higher rates of hypertension (57.0% vs. 41.7%) and diabetes (8.6% vs. 5.0%). For detailed definitions of VRFs, please refer to the **Methods** section. **b**, Illustration of Network Architecture at Nodal/Regional Level. Network integration was assessed using global efficiency (left), which measures the efficiency of parallel information transmission and degree of integration, while segregation was quantified using local efficiency (right) to assess regional resilience to disturbances. The whole-brain architecture is the mean of regional properties. **c**, Distribution of Regional Gender Difference. Throughout the entire brain, males exhibited greater regional network architecture metrics than females. The most significant differences were observed in the medial and superior frontal lobe, superior parietal lobe, medial occipital lobe, and multiple subcortical structures. **d**, Nodal Distribution of BNA-246 Atlas and Gender-Average Networks. A white matter network was constructed for each participant using a finely divided brain template with 246 regions. This panel presents the group-level averaged networks for males and females. **e**, Gender-Specific Aging Trajectories of Whole-Brain Volumetric and Network Measures. GAMs were employed to fit gender-specific aging trajectories for whole-brain network architecture and volumetric metrics, including grey matter volume, white matter volume, and WMH volume. The network architecture exhibited gender differences comparable to other volumetric measures, with males (shown with dashed lines) showing higher whole-brain integration and segregation than females (shown with solid lines) throughout the aging process. The accelerated decline point of whole-brain integration for females occurred around 57.3 years old, which is 9.7 years earlier than males, while the accelerated decline point of whole-brain segregation was approximately 60.5 years old for women and 62.1 years old for men. Abbreviations: M, male; F, female; APOE, Apolipoprotein E; BMI, body mass index; GAM, generalized additive models; ICD, International Classification of Diseases; VRFs, vascular risk factors; WHR, waist-to-hip ratio; WMH, white matter hyperintensity.

### WM architecture is sensitive to gender throughout aging

We assessed the architecture of the WM structural connectomes in 17954 participants by calculating the integration and segregation (Figure 1b) of fiber number-weighted networks (Figure 1d) based on the Brainnetome atlas^18^. The fitted gender-specific aging trajectories of each imaging measures (Figure 1e) indicated that males have greater WM network architecture measures than females throughout the aging process (integration: 7.56±1.22 vs. 6.77±1.06; segregation: 12.4±1.75 vs. 10.9±1.49), and the network architecture was as sensitive as other volumetric measures to gender. The accelerated decline point of the global integration of females appeared about 57.3 years old, which was 9.7 years earlier than that of males, while the accelerated decline point of the global segregation was about 60.5 years old for females and 62.1 years old for males. Computational details are described in the Methods section.

We then investigated the brain regions associated with gender differences in whole-brain network architecture by conducting t-tests for nodal integration and segregation while controlling for age, age², education level, and APOE genotype (Figure 1c). Our analysis revealed that males exhibited greater nodal properties than females throughout the whole brain, with the most prominent differences observed in the medial and superior frontal lobe, the superior parietal lobe, the medial occipital lobe as well as multiple sub-cortical structures. (See **Supplementary Tables 3 and 4** for the nodal gender-difference statistics of the t-tests for integration and segregation, respectively).

### Three key VRFs for whole-brain WM architecture in aging

We separately examined the impact of five VRFs including hypertension, diabetes, excessive alcohol consumption, smoking and obesity on WM network architecture. Considering the pronounced gender differences described earlier, the investigation was carried out separately for two gender groups at the level of the whole brain, brain regions, and functional modules. We conducted t-tests on topological measures that had removed effects from age, age², education, and APOE status. Overall, we identified three key VRFs that were most effective on whole-brain network architecture: diabetes, hypertension, and excessive alcohol consumption (Figure 2a). Smoking and obesity were associated with poorer whole-brain measures in at least one type of architecture in at least one gender group.

**Figure 2.**
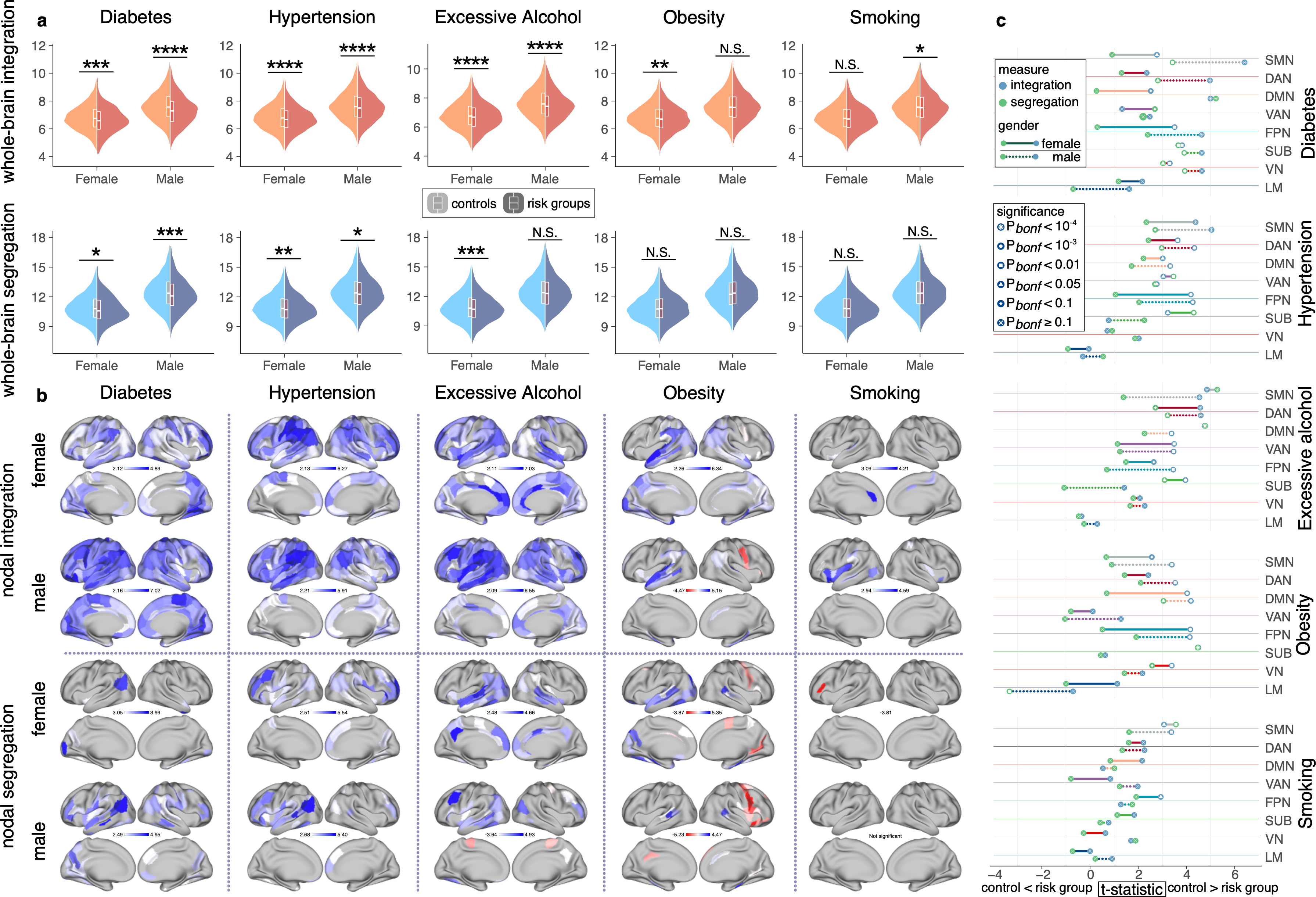
| Single VRF’s Effect on Network Architecture in Each Gender Group. **a**, Effects of Each VRF on Whole-brain architecture. We identified three key VRFs that were most effective on whole-brain network architecture: diabetes, hypertension, and excessive alcohol consumption. In both genders, the presence of any three key VRFs was linked to poorer whole-brain architecture (at a nominal 5% level), except for the effect of excessive alcohol consumption on global segregation in males (male: T = 1.40, p = 0.161; female: T = 4.40, p = 1.1×10^-5^). The influence of obesity and smoking on whole-brain integration varied by sex, with women’s integration being more sensitive to obesity, while men’s whole-brain integration was more affected by smoking. Statistical significance levels are denoted as *: p < 0.05; **: p < 0.01; ***: p < 0.001; ****: p < 0.0001; N.S.: non-significant. **b**, Effects of Each VRF on Regional-brain architecture. We examined the effects of the three key VRFs on nodal integration and segregation in different brain regions. Nodal integration across the entire brain was influenced by the three key VRFs, particularly in the inferior parietal lobule, inferior frontal gyrus, and prefrontal cortex. Nodal segregation exhibited VRF-specific patterns with pronounced sex differences. Female-preferred effects for obesity were observed in the superior temporal gyrus and cingulate gyrus, while male-preferred effects for obesity were observed in the inferior parietal lobule, insular cortex, and occipital gyrus. Regions with significance p-values less than 0.05 after FDR correction are presented in T-maps. **c**, Effects of Each VRF on Modular architecture. Modular integration (blue dots) demonstrated higher vulnerability to VRFs compared to modular segregation (green dots), with males (dashed lines) showing greater vulnerability than females (solid lines). The most pronounced effects were observed in high-order modules, including the DMN, SMN, and attention networks. The VN and SUB were less affected by VRFs, and we note female-specific effects in the SUB to multiple VRFs. No significant effects of any VRF were observed on the architecture of the LM. Abbreviations: FDR, false discovery rate; DMN, default mode network; SMN, somatomotor network; LM, limbic system; VN, visual network; SUB, subcortical network.

Specifically, for whole-brain integration, the presence of any of the three key VRFs was linked with lower integration in both genders, while we observed gender differences of the impact of obesity (male: T = 1.34, p = 0.181; female: T = 2.97, p = 0.0030) and smoking (male: T = 2.53, p = 0.011; female: T = 1.76, p = 0.079). For whole-brain segregation, we only observed significant effects from key VRFs, and we noticed significant effect of excessive alcohol consumption only in males (male: T = 1.40, p = 0.161; female: T = 4.40, p = 1.1×10^-5^). Significance is reported at a nominal 5% level, and please refer to **Supplementary Table 5** for statistics of each VRF’s effect on whole-brain architecture.

### Regional effects of VRFs on WM architecture

We then investigated the differences in nodal topology of 246 brain regions between the risk group and controls for each VRF, separately in male and female groups. The cortical effects are presented as t-maps in Figure 2b, and **Supplementary Table 6** provides detailed statistics of each VRF on nodal measures within each gender group. Generally, VRFs contributed more impairment to nodal integration than segregation, and their effects were more prominent in males than in females except for obesity.

Our investigation into nodal integration has uncovered diffuse effects of three key VRFs across the entire brain, where the inferior parietal lobule, inferior frontal gyrus, and prefrontal cortex exhibited the most prominent effects. Notably, we also have identified a female-specific vulnerability to obesity in multiple regions (Figure 2b). Specifically, the effects of diabetes were found to be diffusive across cortical and subcortical regions in both genders, with males displaying a greater number of significant regions (false discovery rate (FDR) corrected p < 0.05) in the superior frontal gyrus, medial superior temporal gyrus, lateral inferior temporal gyrus, parahippocampal gyrus, lateral superior parietal lobule, dorsal insular cortex, and caudal cingulate gyrus. Hypertension also exhibited diffusive effects in both genders, with the left parietal lobe serving as the center of the effect, while in males, significant regions were identified in the precuneus, right cingulate gyrus, and cuneus. Excessive alcohol consumption diffusely affected cortical and subcortical regions, with the left posterior portions of the brain being the center of the effect. Males displayed a greater number of significant regions in the inferior frontal gyrus, precentral gyrus, inferior temporal gyrus, and postcentral gyrus. Females, on the other hand, displayed a greater number of significant regions in the orbital gyrus, paracentral lobule, superior temporal gyrus, parahippocampal gyrus, and cingulate gyrus. Smoking’s effects were shared among the insular cortex, hippocampus, and thalamus, with males being more susceptible in the inferior parietal lobule, insular cortex, and occipital gyrus, while females were more susceptible in the superior temporal gyrus and cingulate gyrus. The effects of obesity centered in the superior temporal gyrus in both genders, with females displaying greater effects in the middle frontal gyrus, middle temporal gyrus, inferior temporal gyrus, fusiform gyrus, parahippocampal gyrus, superior parietal lobule, inferior parietal lobule, precuneus, insular cortex, cingulate gyrus, occipital gyrus, amygdala, striatum, and thalamus.

We also conducted region-wide analyses on nodal segregation, and we found that the three key VRFs had significant effects in specific brain regions, which exhibited gender-specific patterns. Diabetes was found to primarily affect the left superior parietal lobe, with male-preferred effects in the frontal lobe, temporal lobe, parietal lobule, fusiform, parahippocampal gyrus, precuneus, insular, cuneus, and striatum. Hypertension mainly affects the frontal lobules and angular gyrus, with greater effects on females in the middle frontal gyrus, superior parietal lobule, inferior parietal lobule, and dorsolateral putamen. Excessive alcohol consumption mainly affects the temporal-parietal regions in both genders, with female-specific effects found in several regions, including the superior frontal gyrus, orbital gyrus, parahippocampal gyrus, posterior superior temporal sulcus, inferior parietal lobule, precuneus, insular, cingulate gyrus, thalamus, and striatum. Male-specific effects were found only in the middle frontal gyrus and precentral gyrus. Smoking had only one significant effect that passed the FDR correction, in the left inferior frontal sulcus for females (T = −3.81, p = 0.035). Finally, we emphasize that the protective role of obesity is more prominent in segregation than integration, particularly in the right frontal lobes and precentral gyrus, as people with obesity tended to have altered nodal segregation in several regions, including the orbital gyrus, parahippocampal gyrus, posterior superior temporal sulcus, right frontal lobes, and precentral gyrus, as well as subcortical structures, and female-preferred effects were found in the temporal lobe, fusiform gyrus, precuneus, superior occipital gyrus, amygdala, and thalamus, while male-preferred effects were found in the postcentral gyrus, which has a protective effect, and cingulate. Our findings are important for identifying potential risk factors for cognitive decline and developing personalized intervention strategies to mitigate the effects of VRFs on nodal segregation in the brain.

### High-order functional modules are more susceptible to VRFs

We investigated the module-specific impact of VRFs in seven cortical functional modules and a subcortical module based on Yeo’s classic partition scheme^19^ (**Supplementary Table 7** describes the modules to which each brain region belongs in the Brainnetome atlas). In the modular analysis, Bonferroni correction was applied to account for multiple comparisons across modules, with a threshold of 0.05 for statistically significant corrected p-values. **Supplementary Table 8** provides the statistics of modular comparisons for each gender group.

Our results indicated that VRFs affected modular topology as well with the most pronounced effects observed in high-order modules (Figure 2c, please refer to its caption for a more comprehensive description). The three key VRFs continued to exert a strong influence on modular topology, especially on the integration of high-order networks including somatomotor network, attention network (with greater effects observed in the dorsal part than the ventral part), default mode network and fronto-parietal network. These modules were more susceptible to effects on modular integration than segregation, with males displaying greater susceptibility than females (refer to the top three groups of sticks in Figure 2c).

In contrast, the visual and subcortical modules were less affected by VRFs but exhibited strong female-specific effects in subcortical module (Figure 2c). We found that diabetes influenced the architecture of visual network in both gender groups, whereas obesity only weakly affected visual network in females (For integration, male: T = 2.17, p = 0.24; female: T = 3.39, p = 5.57×10-3; For segregation, male: T = 1.43, p = 0.15; female: T = 2.58, p = 8.00×10-2), and we observed susceptibility of subcortical module to diabetes, and female-specific effects on subcortical module from excessive alcohol consumption (For integration, male: T = 1.42, p = 0.16; female: T = 3.96, p= 6.09×10-4; For segregation, male: T = −1.07, p = 0.28; female: T = 3.09, = 1.60×10-2), hypertension (For integration, male: T = 0.77, p = 0.44; female: T = 3.23, p = 1.00×10-2; For segregation, male: T = 2.26, p = 0.193; female: T = 4.30, p = 1.36×10-4) and obesity (For integration, male: T = 0.63, p = 0.53; female: T = 4.50, p = 5.70×10-5; For segregation, male: T = 0.44, p = 0.66; female: T = 4.47, p = 6.49×10-5) . We did not observe significant effects of any VRF on the architecture of the limbic system.

### Vascular risks have greater effects on WM architecture in males

We employed a structural equation modeling approach to examine the directed associations between various factors, including vascular risk factors, WM hyperintensity volume, WM network architecture, age, and cognitive test performance. The results were analyzed separately for the global integration and segregation of females and males, as depicted in Figure 3a and b. All pathways in the four models displayed in Figure 3a and b were found to be statistically significant (p<0.05), with the exception of the relationship between excessive alcohol consumption and general vascular risk in females (model of integration: β = 0.27, p = 0.076 ; model of segregation: β = 0.06, p = 0.121), the impact of WM hyperintensity volume on global segregation in females (β = −0.05, p = 0.051), the impact of vascular risk on global segregation in females (β = −0.046, p = 0.217), and the effect of WM hyperintensity volume on general cognition in males in the model of integration (β = −0.05, p = 0.068). All models in Figure 3a and b exhibited root mean square error of approximation values less than 0.08, indicating a good fit with the data.

**Figure 3.**
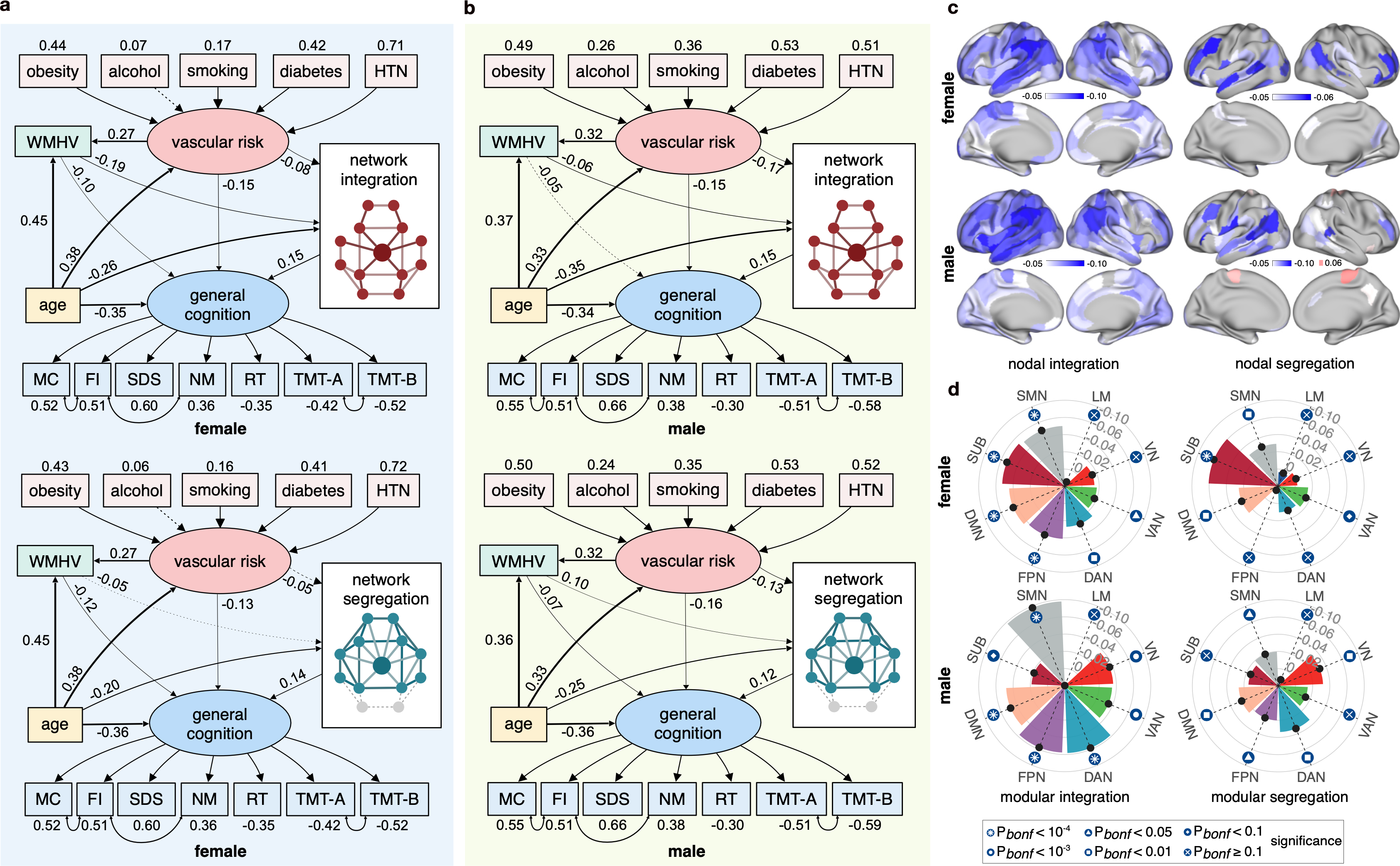
| Structural equation model and the effects of combined risk on network architecture. **a, b** Full Structural Model in Females **(a)** and Males **(b)**. Standardised beta coefficients are presented. All pathways in the four models were statistically significant (p<0.05), with the exception of the relationship between excessive alcohol consumption and general vascular risk in women (model of integration: β=0.27, p =0.076 ; model of segregation: β=0.06, p =0.121), the impact of WMHV on global segregation in women (β=−0.05, p=0.051), and the effect of WMHV on general cognition in men (model of integration: β=−0.05, p =0.068 ; model of segregation: β=−0.07, p =0.014). **c**, Effects of CRS on Nodal Architecture. We computed CRS for each participant based on the coefficients obtained from the structural equation models (See **Methods** section for details). The effects on nodal integration were primarily concentrated in the bilateral inferior parietal cortex, left temporal lobe, and left frontal lobe. The impact on nodal segregation was mainly observed in the bilateral prefrontal cortex and inferior parietal cortex. Gender differences were observed, with a more pronounced effect on nodal properties in the temporo-parietal region in males compared to females, and weaker effects in the lateral prefrontal cortex in males. The β-maps after FDR correction are presented. **d**, Effects of CRS on Modular Architecture: CRS had significant effects on all modules except the limbic system (Bonferroni corrected p < 0.05). Gender differences were found in the VN and SUB, with the integration and segregation of VN unaffected by CRS in females, and the architecture of SUB unaffected in males. Abbreviations: CRS, Combined Vascular Risk Score; FDR, False Discovery Rate; SUB, Subcortical; VN, Visual Network; WMHV, White Matter Hyperintensity Volume.

Elevated general vascular risk correlated with worse WM network architecture. Notably, the pathway of WM network impairment shows gender specificity with WM network impairment derived from vascular risk is more mediated by WM hyperintensity volume in females while males show more WM network impairment directly from vascular risk compared to the part mediated by WM hyperintensity volume. However, we didn’t see great gender disparity in the impact of age, vascular risk, WM hyperintensity volume, and network architecture on general cognition.

Based on the path weights in Figure 3a and b, we further calculated the combined vascular risk score (CRS) and examined the impact of CRS on the nodal and modular architecture while controlling age, age², education level, and APOE status within each gender group (Figure 3c and d, see **Supplementary Table 9** for nodal statistics and **Supplementary Table 10** for modular statistics). Our analysis revealed that the effects of CRS on the nodal integration was primarily concentrated in the bilateral inferior parietal cortex and the left temporal lobe and left frontal lobe, while the impact on nodal segregation was mainly observed in the bilateral prefrontal cortex and inferior parietal cortex (Figure 3c). Notably, we observed gender differences, with a more pronounced effect on nodal properties in the temporo-parietal region in males compared to females, and weaker effects in the lateral prefrontal cortex in males. At the modular level (Figure 3d), CRS had significant effects all modules except the limbic system (Bonferroni p < 0.05). We also found gender differences in visual and subcortical modules, with the integration (male: β = −0.056, p = 5.5×10^-5^; female: β = −0.035, p = 1.59×10^-2^) and segregation (male: β = −0.052, p= 2.2×10^-4^; female: β = −0.023, p = 0.12) of visual network unaffected by CRS in females, and the architecture of subcortical modules unaffected in males (for integration male: β = −0.039, p = 6.9×10^-3^; female: β = −0.073, p = 4.7×10^-7^; for segregation male: β = −0.035, p = 1.52×10^-2^; female: β = −0.081, p = 2.5×10^-8^).

### Combined vascular risk affects processing speed and working memory

We further evaluated the effect of CRS on each specific neuropsychological test performance in participants independent of brain-network construction. Results indicate significant, yet weak, effects of CRS on reaction time (β = 0.046, p = 5.18×10^-4^, N = 12475), numeric memory (β = −0.031, p = 0.027, N = 12056), symbol digit substitution (β = −0.052, p = 5.43×10^-5^, N = 11991), and matrix pattern completion (β = −0.034, p = 0.012, N = 12055), while no significant effects were observed on fluid intelligence (p = 0.817, N = 12584), pairs matching (p = 0.340, N = 12559), TMT-A (p = 0.073, N = 11991), TMT-B (p = 0.957, N = 11985), and tower rearrange (p = 0.232, N = 11979). These findings suggested that combined vascular risk had a greater impact on processing speed, working memory and visual-spatial memory than on reasoning and executive function (See **Supplementary Table 11** for detailed statistics).

## Discussion

### Gender-common effects of vascular risk factors

In this study, we investigated the influence VRFs on the aging WM network architecture in separate gender groups. Consistent with previous research, hypertension^7^, diabetes^20^, and excessive alcohol consumption^9^ emerged as the most influential factors negatively affecting the WM network architecture in both genders, particularly on network integration. The inferior parietal lobule, inferior frontal gyrus, and prefrontal cortex showed the greatest vulnerability to these VRFs. The effective patterning aligned with prior study linking high blood pressure to compromised global WM network integrity, primarily associated with disconnections in frontal and parietal regions^7^. However, our findings pertaining to diabetes and alcohol overconsumption exhibited discrepancies with prior research, likely due to variations in sample characteristics across different studies^7, 8, 20^. We note that the nodal disruption of excessive alcohol consumption on frontal and temporal regions provides a potential neural basis of attenuated functional responses in the right fronto-temporal cortex linked to alcohol-induced impairment of inhibitory control^21^. Overall, our study sheds light on the influence of VRFs on the aging WM network architecture, emphasizing the importance of implementing interventions aimed at controlling hypertension, diabetes, and excessive alcohol consumption as a priority.

The effect sizes observed in our study were predominantly small, particularly when examining the impact of smoking and obesity on brain health. Consistent with previous research^11^, obesity does not exert substantial brain-wide effects on network architecture in either sex group. However, it is essential to acknowledge that our observations may have been influenced by thresholding effects, as a previous UKB-based study have identified a linear correlation between body mass index, waist-to-hip ratio, and bilateral temporal atrophy^22^. Furthermore, our study highlights the significant influence of smoking on the integration of the insula and the rostral hippocampus in both genders. Considering that individuals with insula damage^23^ or reduced insula-related functional connectivity^24^ show decreased reliance on substances like cocaine, the abnormal increase in functional connectivity associated with smoking may serve as a compensatory mechanism to counterbalance the structural decline of insula integration. Despite the relatively small magnitudes in effect sizes, our results offer important insights into the relevance to brain health.

We further investigated the impact of CRS on the architecture of functional modules and cognitive functions. The most significant effects were observed on the SMN and the FPN, as well as on working memory and reaction speed. Regional analyses suggest that CRS mainly affects the topological efficiency in areas with high metabolism^25^, which underlie the core of high-order cognitive functions, such as the striatum, thalamus, and cortical hubs^2^. Our study reinforces the assumption rising from tract-based studies^22^, that VRFs disrupt WM fiber tracts connecting key brain regions, leading to structural disconnection, atrophy, and cognitive dysfunction. The impairment of the dorsolateral prefrontal cortex, which regulates working speed and ensures accurate performance and serves as the core area of FPN^26^, indicates that VRF-related deterioration of white matter topology supporting attention resources, coordination, and accuracy explains the decrease in reaction speed. These findings offer valuable insights for developing targeted interventions that aim to mitigate cognitive decline by addressing combined vascular risks, thereby promoting brain health, and potentially enhancing cognitive function in a more complex scenario.

### Gender-related effects of vascular risk factors

Our study presents a pioneering investigation of gender differences in the macro-scale WM network architecture during the aging process. Remarkably, we demonstrate that women exhibit consistently lower network architecture throughout aging span and experience an earlier onset of accelerated decline compared to men. However, we note that despite differences in brain volume and network architecture between genders, the contribution of network architecture, aging and vascular risk factors on general cognition remained similar between two gender groups (Figure 3). We also note the distribution of regional architectural difference highly resemble the DMN that is key in the spectrum of multiple neurodegenerative diseases, which may explain the neural basis of a higher prevalence of female^14^. Multiple biological factors, including variations in estrogen levels^27^, chronic inflammation^28^, and genetic influences^29^, likely contribute to the intricate etiology underlying the observed network architecture changes and gender-differences during aging. Further investigations are warranted to comprehensively elucidate the complex interplay of these factors and their implications for gender disparities in the aging brain.

We have observed gender-related discrepancies in the impact of risk factors on network architecture, specifically in relation to diabetes and obesity. Notably, we found a male-specific effect of diabetes on integration within the SFG, while obesity demonstrated a female-specific and multi-regional influence. Our study revealed that obesity-related integration disruptions were predominantly observed in females including the thalamus, striatum, amygdala, left occipital lobe, left MTL, and SPL. Interestingly, one previous research has indicated that increased BMI among aging females is associated with heightened connectivity in the bilateral frontoparietal and parahippocampal regions of the cingulum^12^. Building upon this, our findings suggest a protective effect of obesity on the structural connectivity of the left MTL in women, potentially indicating a compensatory role of the MTL in response to long-term inflammation associated with obesity^30^. Future studies identifying gender-specific disparities of how diabetes and obesity impact neural connectivity will help develop tailored strategies for both genders.

### Limitations and future directions

Our study has certain limitations that warrant careful consideration in the interpretation of our findings. Firstly, our study participants were confined to individuals aged 50 to 80 years. While our investigation provides insights into the effects of VRFs on the brain within this age group, it is important to acknowledge that the impact of these factors on cerebral vascular structure and white matter lesions in younger populations has been extensively studied^31^. Thus, our study may not offer a comprehensive understanding of the effects on younger individuals, necessitating caution in generalizing the findings to broader age ranges. Secondly, the participants in our study may not be fully representative of the general aging population in the UK. The selection of individuals from the UK Biobank as our study cohort introduces the possibility of a bias toward individuals with relatively favorable living conditions. Consequently, the generalizability of our results to wider populations with diverse socioeconomic backgrounds and living conditions may be limited. Thirdly, the study design relied on cross-sectional data obtained from the UK Biobank, thereby limiting our ability to establish causal relationships. Although cross-sectional studies provide valuable insights into associations between variables, they cannot ascertain the direction of causality. To elucidate the temporal relationships between vascular risk factors and brain health, future longitudinal studies are necessary. Moreover, the brain parcellations employed in our study were derived solely from the cortical and subcortical regions of cerebrum, which restricts the capture of aging-related patterns in other regions of the central nervous system. Lastly, the cognitive tests administered in the UK Biobank were conducted using touch-screen devices, albeit under the supervision of a nurse. While efforts were made to standardize the administration process, it is important to recognize that this method may not fully align with the clinical standards typically employed in cognitive assessments. Considering these limitations, it is imperative to interpret the findings of our study with caution and recognize the potential necessity for further research to explore these relationships across different age groups, populations, and assessment settings.

In our study, we utilized binarized metrics unanimously for all VRFs instead of continuous metrics. We made this decision based on two main considerations. Firstly, the quality of the available data did not allow for a fully precise and dynamic description of the VRF status. Secondly, we lacked precise parametric thresholds for each VRF on network health, particularly in the case of smoking. As a result, the potential non-linear relationship between these VRFs and the network architecture remains unknown. To address this gap in knowledge, future research should explore whether VRFs affect white matter network architecture in a dose-dependent or parametric manner^32^. Deriving precise thresholds or parametric relationships for each VRF based on network architecture can gain a more nuanced understanding of the association between VRFs and white matter network architecture. We also note it is crucial for future studies to have a comprehensive design that takes into account the strong interplay among various risk factors such as the comorbidity rate of hypertension and diabetes^33^, which would be important in order to disentangle the specific contributions of each risk factor to the observed effects on white matter network architecture.

## Conclusions

Our study investigated the impact of VRFs and gender on aging white matter network architecture using brain imaging data from 17,954 participants in the UK Biobank. We found that females exhibited lower white matter network architecture compared to males, as well as an earlier degeneration of the network architecture. Higher vascular risk was associated with poorer white matter network architecture and specific cognitive impairments, particularly in processing speed and working memory. Hypertension, diabetes, and excessive alcohol consumption were identified as three key VRFs causing the most damage to the white matter network architecture, with the lateral temporal lobe being the primary site of damage. Our findings contribute to understanding the relationship between VRFs, gender, and aging white matter network architecture, providing insights into the underlying mechanisms of white matter degeneration and its implications for cognitive function in aging populations. Further research is warranted to develop targeted interventions addressing the impact of VRFs on brain health and cognitive decline.

## Methods

### Participants

From the UK Biobank data release, we selected 20007 participants who had undergone the imaging visit since 2014 with available structural and diffusion MRI scans and questionnaire survey information on lifestyle and medical history. During the MRI scan assessment, participants were asked to provide demographic, health, and socioeconomic information via touchscreen responses. To enhance the accuracy of the data, nurses inquired about participants’ medical histories, including any self-reported diagnoses. Participants with a history of chronic degenerative neurological disorders (including demyelinating disorders), brain cancer, cerebral hemorrhage, brain abscess, aneurysm, cerebral palsy, encephalitis, head injury, nervous system infection, head or nerve injury or trauma, or neuropsychiatric disorders were excluded. We performed further exclusion with WM structural networks constructed with the structural and diffusion-weighted MRI for participants who passed the screening. We excluded 2053 participants based on the quality control of atlas transformation and tracking plausibility (See Constructing WM connectomes), resulting in 17,954 participants with structural brain network. Written and informed consent was obtained from all participants, and the study was approved by the Research Ethics Committee (REC number: 11/NW/0382). **Supplementary Text 1** provides the ICD-10 codes as well as related data-field of self-report for disease exclusion. **Supplementary Table 2** provides information on the missing numbers for each neuroimaging measure, medical measure, and neuropsychological tests.

### Vascular risk factors

We considered five VRFs commonly studied in the literature for their significant negative impact on the central nervous system^5, 6, 34^. These included two vascular disease risk factors, hypertension, and diabetes, as well as three lifestyle risk factors, smoking, excessive alcohol consumption, and obesity. Diabetic participants met any of the following conditions: once diagnosed with diabetes, meeting the definition of diabetes in ICD-10, taking insulin, blood sugar level ≥ 7 mmol/L, or glycosylated hemoglobin level ≥ 48 mmol/mol. Hypertensive participants met any of the following conditions: once diagnosed with hypertension, meeting ICD-10 criteria for high definition of blood pressure, taking blood pressure drugs, systolic blood pressure ≥ 140 mmHg, or diastolic blood pressure ≥ 90 mmHg. Excessive drinking participants were considered to have both a history of drinking and an average daily alcohol intake of not less than 14 grams for females or 28 grams for males according to US dietary guidelines^35^. Smokers were defined as individuals who reported ever smoking or pack-years greater than 0. Obese participants were defined as individuals with a body mass index ≥ 30 or a waist-to-hip ratio greater than 0.85 for females or 0.9 for males. For detailed definitions of each vascular risk factor and the associated UK Biobank data filed codes, refer to **Supplementary Table 1** and online codes.

### Neuropsychological testing

The UK Biobank carried out large-scale neuropsychological testing without the presence of an examiner. Details on the UK Biobank neuropsychological tests have been described elsewhere and we selected seven tests with good test-retest reliability from these tests^36, 37^. Briefly, the Reaction Time test measured the speed of responses to visual stimuli, and we used the mean time taken to correctly identify matches in milliseconds (data field = 20023). The Numeric Memory test evaluated participants’ ability to remember strings of numbers of increasing length over a short period. We used the maximum number of digits correctly remembered (data field = 4282) as a performance indicator on this test. The Fluid Intelligence test assessed the capacity to solve numerical and verbal reasoning problems, in which participants were given 13 questions to answer in two minutes, and we considered the total number of correct answers given (data field = 20016). The Trail Making Test, where participants were presented with a set of numbers or letters scattered on the screen and were asked to click on them sequentially following a specific algorithm, consisted of two sets of tests.

We used the measure of the time taken to complete the numeric path (TMT-A, data field = 6348) and the alphanumeric path (TMT-B, data field = 6350). The Matrix Pattern Completion test presented participants with a series of matrix pattern blocks with missing elements, and then asked them to choose from a series of displayed options the element that best completed the pattern. The number of puzzles solved correctly (data field = 6373) was used as an indicator of performance on this test. The Symbol Digit Substitution test required participants to use a grid to locate the number associated with each symbol, and we used the number of correctly matched symbol-digit pairs (data field = 23324).

### MRI acquisitions and processing

In this study, three modalities of MRI scans were used, including T1-weighted MRI, T2-weighted MRI, and diffusion-weighted MRI. Implementation standards of neuroimaging acquisition and processing pipeline have already been well scripted elsewhere^38^ (also see Code availability). T1-weighted MRI was acquired with a 3D magnetization-prepared rapid gradient echo sequence with sagittal scan orientation, while T2-weighted MRI was obtained using a 3D fluid-attenuated inversion recovery sequence, both with a voxel size of 1×1×1 mm and in-plane acceleration factor of 2. The diffusion-weighted MRI scan utilized an echo-planar imaging sequence, a voxel size of 2.0×2.0×2.0 mm, and partial Fourier sampling factors of 6/8. The diffusion-sensitizing gradients included 10 b = 0 s/mm² volumes, 50 b = 1 000 s/mm² volumes, and 50 b = 2000 s/mm²volumes, with an echo time of 92 ms and a repetition time of 3600 ms.

UK Biobank performed pre-processing on MRI data with FMRIB Software Library^39^ (see Code availability). T1-MRI underwent gradient distortion correction, followed by cropping to optimize the field of view. Subsequently, brain scalping was performed to remove cranial matter, and FMRIB’s Automated Segmentation Tool^40^ was used to obtain the bias field and components of gray matter, white matter, and cerebrospinal fluid. The volume of each component was then determined.

For T2-weighted MRI, gradient distortion correction was performed prior to linear transformation to T1 space and bias field correction. The corrected image, along with the white matter mask and manually delineated lesion images, were inputted into the Brain Intensity Abnormality Classification Algorithm program^41^. This generated a probability map of WM hyperintensities, which was then binarized using a threshold of 0.8 to determine the distribution and thus enable the calculation of WM hyperintensity volume.

For diffusion-weighted MRI, preprocessing involved eddy current correction, head movement correction, and gradient distortion correction of the original image in combination with a field map estimated from posterior-to-anterior and anterior-to-posterior directed non-diffusional scans. The preprocessed image was then analyzed using the bedpostx tool, which employs model-based spherical deconvolution to estimate up to 3 fiber orientations per voxel. This approach allowed for within-voxel modeling of multi-fiber tract orientation structure, using Bayesian Estimation of Diffusion Parameters Obtained using Sampling Techniques^42^.

### Constructing WM connectomes

To construct the WM connectome, we registered the fine Brainnetome Atlas with 246 parcellations defined in standard Montreal Neuroimaging Institute space^18^ and the WM mask in the T1 space to each participant’s naive diffusion space. We used Camino (http://camino.cs.ucl.ac.uk/) to perform deterministic WM fiber tracking on the ball-stick model in the diffusion space, with the transformed WM mask used as seeding regions^43^. We utilized nearest-neighbor interpolation and the Fourth-order Runge-Kutta method as tracking algorithm, with the tracking step size set to 2mm. To maintain biological plausibility, we discarded compartments with a mean volume fraction below 0.1 and terminated tracking if curvature exceeded 45 degrees at each 5 mm interval. We further post-processed the tracking results by removing fibers with a length below 20mm or above 250mm in the individual diffusion space. Finally, we used the conmat command to construct a fiber number-weighted connectivity network based on the Brainnetome atlas. To ensure accuracy, we had five experienced colleagues visually cross-check the transformations of the Brainnetome atlas and the plausibility of tracking. From the 20,007 participants with downloaded diffusion-weighted and structural scans, we excluded 2,015 participants whose image failed the pipeline and 38 participants due to poor results. For a detailed description of the construction, please refer to the online code (see Code availability).

### Quantifying network architecture

On the construction of structural networks, we assessed the architecture of each participant’s WM structural network in terms of integration and segregation. Global efficiency is usually considered as a measure of the efficiency of parallel information transmission and the degree of integration (Figure 1b, left)^44^. For a given node *i* within a network *N* consisting of n nodes, the nodal global efficiency of *i* was defined as:

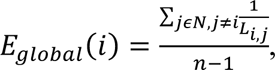

where the *L_i,j_* represents the shortest path length between node *i* and node *j* in network *N*.

The integration of network *N* was then defined as the mean nodal global integration across the system, as calculated by:

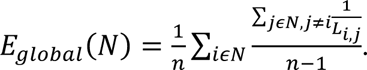

In addition to global efficiency, nodal efficiency was used to quantify the segregation and resilience to disturbances^45^. The local efficiency of node *i* was determined by the global integration of the subnetwork *N_i_*, which consisted of *i*’s neighbors with the connections among them, i.e. *_local_*(*i*) = *_glocal_*(*N_i_*) (Figure 1b, right). The segregation of network *N* was computed as the mean segregation across all nodes by:

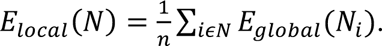

We further extracted seven cortical modules and one subcortical module from the whole brain network of each participant according to Yeo’s functional partitioning scheme^19^. To perform the modular analyses (see Statistical methods), we used the extracted modules as input to calculate the modular integration and separation. **Supplementary Table 8** described the Yeo’s partitioning scheme based on the Brainnetome atlas. All graph theory calculations were performed using the Gretna toolbox^46^.

### Statistical methods

To examine the sensitivity of the architecture of the network to gender and aging compared with volumetric measures, we employed generalized additive models^47^ to fit WM network architecture and volumetric measures by age in different gender groups. Furthermore, we calculated the accelerated declining points of each measure during the aging process by calculated the second-order difference of the fitting curves. We used chi-square tests to compare discrete demographic information data by gender and t-test to compares continuous demographic information data by gender.

To explore the effects of VRFs on WM network architecture in both genders, we conducted two-sided t-tests between the risk group and controls on the residuals removing the effects of age, age², education level and APOE genotype. These analyses were performed at the regional, modular, and whole-brain levels. To ensure robust control for false positives, we corrected p-values using FDR correction for region-wide statistical results and Bonferroni correction for module-wide results.

Finally, we conducted the structural equation modelling analysis separately in males and females to investigate the directed relationships among age, multiple VRFs, WM hyperintensity volume, brain network architecture and cognition. We further calculated CRS for each participant *i* based on the path weights to ‘vascular risk’ in structural equation models (See Figure 3a and 3b), as defined by:

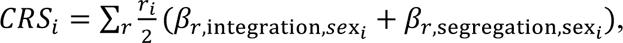

where *r* is a risk factor with dichotomous value among five VRFs and *β* is the standardized coefficient in structural equation models.

We validated the plausibility of CRS with linear models examining the effects of CRS on network architecture of the two gender groups at the regional, modular, and whole-brain level. Age, age², education, and APOE genotype were included in linear models as covariates. Again, we modeled each brain region and module, and used FDR and Bonferroni correction for statistical significance at the region and module level, respectively. Additionally, we performed linear regressions of CRS on each cognitive performance in an independent sample of WM network construction, controlling for the above covariates. See Figure 3a and 3b for the structural equation models.

Statistical analyses were conducted using R 4.1.0, except for structural equation model analyses, which were performed using MPlus^48^ (see Code availability). Outliers were removed from all models, whereby outliers were defined as data points whose values of the response variable deviated from the mean by more than three standard deviations. Brain regions that exhibited statistical significance after FDR correction (FDR corrected p-value < 0.05) were visually represented with t-statistics, while we adopted a Bonferroni-corrected p-value threshold of 0.05 for module-wide statistics. We used BrainNetViewer Toolbox^49^ to visualize the brain network and Workbench software^50^ to visualize regional statistics.

## Data Availability

All data produced in the present study are available upon reasonable request. All data produced are available online at

https://biobank.ndph.ox.ac.uk/

